# Machine learning-optimized Combinatorial MRI scale (COMRISv2) correlates highly with cognitive and physical disability scales in Multiple Sclerosis patients

**DOI:** 10.1101/2021.03.26.21254405

**Authors:** Erin Kelly, Mihael Varosanec, Peter Kosa, Mary Sandford, Vesna Prchkovska, David Moreno-Dominguez, Bibiana Bielekova

**Affiliations:** Neuroimmunological Diseases Section, Laboratory of Clinical Immunology and Microbiology, National Institute of Allergy and Infectious Diseases, National Institutes of Health, Bethesda, MD; QMENTA, Boston, USA

## Abstract

Composite MRI scales of central nervous system tissue destruction correlate stronger with clinical outcomes than their individual components in multiple sclerosis (MS) patients. Using machine learning (ML), we previously developed Combinatorial MRI scale (COMRISv1) solely from semi-quantitative (semi-qMRI) biomarkers. Here, we asked how much better COMRISv2 might become with the inclusion of quantitative (qMRI) volumetric features and employment of more powerful ML algorithm.

The prospectively acquired MS patients, divided into training (n=172) and validation (n=83) cohorts underwent brain MRI imaging and clinical evaluation. Neurological examination was transcribed to NeurEx app that automatically computes disability scales. qMRI features were computed by LesionTOADS algorithm. Modified random forest pipeline selected biomarkers for optimal model(s) in the training cohort.

COMRISv2 models validated moderate correlation with cognitive disability (Rho = 0.674; Linh’s concordance coefficient [CCC] = 0.458; p<0.001) and strong correlations with physical disability (Spearman Rho = 0.830-0.852; CCC = 0.789-0.823; p<0.001). The NeurEx led to the strongest COMRISv2 model. Addition of qMRI features enhanced performance only of cognitive disability model, likely because semi-qMRI biomarkers measure infratentorial injury with greater accuracy.

COMRISv2 models predict most granular clinical scales in MS with remarkable criterion validity, expanding scientific utilization of cohorts with missing clinical data.

## Introduction

Structural imaging of the central nervous system (CNS) by magnetic resonance imaging (MRI) plays central role in diagnosing multiple sclerosis (MS) and evaluating efficacy of treatments. Nevertheless, the correlations between any MRI biomarker and clinical disability measures are only mild to moderate.

This is explainable by following shortcomings of both clinical scales and MRI biomarkers: A. Reliability: this includes technical aspects of the measurement such as test-retest variability, different scanners or different analysis methods; variability between different raters, and B. Criterion validity: this refers to how each measurement reflects true CNS tissue damage.

While original description of most expert-derived clinical scales missed test-retest reliability (e.g. Expanded Disability Status Scale [EDSS])^1^, the clinical trials identified “transient worsening and improvements” in approximately 20% of subjects. Understanding that these fluctuations likely represent measurement noise, recent trials “confirmed” disability progression at 3-6 months of follow-up. We assessed inter-rater variability of neuroimmunology clinical scales, not by examining identical patient(s) by different clinicians, but by transcribing the same neurological examination documented in the structured electronic medical record note into 4 clinical scales.^2^ Although when comparing hundreds patient/exams, the concordance coefficients (CCC; measure that reflects 1:1 concordance of the ratings, with 0 representing no concordance and 1 representing 100% concordance) between two clinicians were excellent (i.e., ranging from 0.943-0.968; p-value < 1×10^−7^), the difference for a single exam represented up to 3 EDSS points. Therefore, we developed NeurEx iPad app that algorithmically codifies translation of a neurological examination into 4 clinical scales after a clinician rapidly documents neurological deficits using highly intuitive touch screen interphase on homunculus icons ^2^. By eliminating variance from one of the clinicians (i.e., NeurEx always provides only one rating per clinical scale for a given neurological exam), we increased inter-rater reliability between single clinician and NeurEx app to a maximum difference of 1.5 EDSS points (CCC 0.968-0.987; p-value < 1×10^−7^). Of course, NeurEx can’t eliminate noise stemming from variances in the performance of neurological examination by different clinicians and this likely represents the greater source of noise.

Even more pressing limitation of traditional clinical scales is their sensitivity and construct/criterion validity. For example, natural history cohorts show that average MS patient progresses by 1 EDSS point every 10 years ^3, 4^. Clearly, many axons demyelinate, and oligodendrocytes/neurons die during those 10 years and this ongoing CNS tissue destruction is not captured by EDSS. Additionally, while large areas of CNS tissue (especially brain, but also cerebellum) serve cognition, our ability to reliably quantify complex cognitive functions is extremely limited. Consequently, cognitive functions are severely underrepresented in traditional disability scales. A creative attempt to remedy these limitations was MS functional composite (MSFC), an expert-derived composite scale of 3 functional tests of equal weight, reflecting ambulation, fine finger movements and memory/processing speed ^5^. While concept of MSFC was outstanding, one of the selected components, Paced Auditory Serial Addition Test (PASAT3) proved suboptimal, suffering from high test-retest variability and a learning effect.

Avoiding mistakes in MSFC development, we used data-driven (i.e., machine learning [ML]) approach to select contributing features and their optimal “weights” to derive Combinatorial, weight-adjusted Disability Score (CombiWISE; continuous scale from 0-100) ^6^. Unfortunately, neither PASAT3 nor alternative cognitive test Symbol Digit Modalities Test (SDMT; ^7^) were selected by ML. In two independent validation cohorts CombiWISE correlated strongly with EDSS and measured progression of disability in time intervals as short as 6-12 months. Nevertheless, even this granular clinical scale undoubtedly lacks sensitivity to measure daily destruction of individual axons/neurons and oligodendrocytes, likely happening in MS.

The insensitivity of clinical scales to underlying cellular events is unsurmountable, as thousands of neurons and large synaptic circuits underly measurable neurological functions, and functional repair (i.e., remyelination and new synapse formation by remaining neurons) masks clinical deficit. Thus, MRI-based structural imaging and quantification of cellular substructures using advanced imaging methods such as magnetization transfer imaging (MTR) or diffusion tensor imaging (DTI) raised hopes for objective measurements of CNS tissue destruction that may outperform sensitivity of clinical scales.

Unfortunately, MRI biomarkers proved to have their own limitations. Seeing the exquisite details of CNS tissue captured by MRI, we forget that MRI does not measure tissue structure directly. MRI deduces CNS structure from the decay of energized hydrogen protons, which is highly dependent on the technical aspects of specific MRI machine and acquisition protocols, on complex post-processing algorithms but also on transient biological processes such as subjects’ hydration, use of alcohol or pharmaceutical agents ^8^. Consequently, test-retest variability of MRI biomarkers, compounded by movement artifacts in living human subjects, is high in comparison to measured change. This leads to poor signal-to-noise ratio (SNR), except for semi-quantitative MRI features (semi-qMRI) such as contrast-enhancing lesions or distinct T2 lesions formed in different CNS compartments ^6, 9, 10^.

Additionally, the criterion validity of any single MRI biomarker is problematic as all capture only some aspects of MS-related CNS tissue destruction and do so with restricted specificity. To surpass this limitation, several groups explored combinations of MRI features, akin to composite clinical scales ^11-17^. All published combinatorial MRI models outperformed each contributing MRI biomarker in correlation with clinical outcomes, validating this concept.

Like in combinatorial clinical scales, most groups selected contributing MRI features based on expert opinions and persisted in these choices even when unilateral correlations proved alternative MRI feature(s) superior ^18-21^. Instead, we took data driven approach to develop COMRISv1 (Combinatorial MRI scale, version 1), where both selection of contributing features and their weights in the final model were derived from unbiased ML approach ^22^. In contrast to other combinatorial MRI scales, COMRISv1 was derived from semi-qMRI, instead of quantitative MRI (qMRI) features to improve SNR, while, inevitably, sacrificing sensitivity. Despite this, when tested in the independent validation cohort, COMRISv1 models achieved the highest correlations with physical (i.e., EDSS; Rho = 0.7, p-value<0.001, n=114) and cognitive (i.e., SDMT; Rho = 0.5, p-value<0.0001, n=92) disability among all published combinatorial MRI scales for MS.

Nevertheless, we wondered, and this paper answers, whether incorporating volumetric qMRI features and using more powerful ML models would strengthen performance of COMRISv2.

## Methods

### Subjects, ethics approval and consent to participate

All subjects were prospectively recruited to the National Institute of Allergy and Infectious Diseases (NIAID) of the National Institutes of Health (NIH) natural history protocol “Comprehensive Multimodal Analysis of patients with Neuroimmunological Diseases of the CNS”; Clinicaltrials.gov identifier NCT00794352. The study was approved by NIAID scientific review and by the NIH Institutional Review Board. All methods were performed in accordance with the relevant guidelines and regulations. All subjects provided written informed consent. Table 1 contains demographic and clinical data on all subjects.

**Table 1:** Demographic data for training and validation cohort. (RRMS – Relapsing-Remitting Multiple Sclerosis, PPMS – Primary Progressive Multiple Sclerosis, SPMS – Secondary Progressive Multiple Sclerosis, CIS – Clinically Isolated Syndrome, RIS – Radiologically Isolated Syndrome, SDMT – Symbol-Digit Modalities Test, EDSS – Expanded Disability Status Scale, SNRS – Scripps Neurological Rating Scale, CombiWISE – Combinatorial Weight-adjusted Disability Scal, SD – standard deviation).

|  | *Discovery cohort* | *Validation cohort* |
| --- | --- | --- |
| **N (% Female)** | 172 (52.9) | 83 (59.0) |
| **% RRMS/SPMS/PPMS/CIS-RIS** | 41.3/22.1/33.7/2.9 | 34.9/28.9/31.3/4.8 |
| **Age (mean, SD)** | 53.2 (12.3) | 53.0 (12.1) |
| **SDMT (mean, SD)** | 44.1 (14.2) | 42.7 (13.7) |
| **EDSS (mean, SD)** | 4.8 (1.9) | 4.8 (1.9) |
| **SNRS (mean, SD)** | 64.9 (16.4) | 64.5 (14.8) |
| **CombiWISE (mean, SD)** | 39.4 (17.6) | 39.5 (16.5) |
| **NeurEx (mean, SD)** | 127.7 (93.4) | 126.1 (80.5) |

### Collection and computation of clinical scales

All participants underwent a comprehensive diagnostic process, including neurological examination transcribed to iPad-based app NeurEx, which automatically calculates several clinical scales, including Expanded Disability Status Scale (EDSS; ordinal scale from 0-10) and streams data to Neuroimmunological Diseases Section (NDS) research database hosted on secured NIAID server.

Another set of investigators, blinded to NeurEx data collected timed 25-foot walk (25FW), 9-hole peg test (9HPT) and SDMT and inputted these to the NDS database. The database automatically integrates data to calculate the Combinatorial Weight-Adjusted Disability Score (CombiWISE; continuous scale from 0-100). NDS database has also user-defined privileges that blind the clinicians and investigators collecting clinical and functional data to qMRI and semi-qMRI data. MS diagnosis was based on 2010 McDonald diagnostic criteria ^23^ and, after 2017, based on its 2017 modifications ^24^.

### Collection and computation of MRI biomarkers

Brain MRIs were performed on Signa – (1.5T and 3 Tesla, General Electric, Milwaukee, WI) and Skyra – (3 Tesla, Siemens, Malvern, PA) Units using 16 – and 32 – channel imaging coils with previously-described scanning protocols ^22^. Our brain MRI sagittal and axial cuts extend distally to C5 level, allowing determination of semi-qMRI biomarkers of medulla/upper spinal cord (SC) atrophy and lesion load.

The semi-qMRI data were acquired by consensus of MS-trained clinicians during weekly clinical care meetings. The rating of semi-qMRI features was previously extensively described ^22^ and its codification was integrated to NDS research database.

To acquire qMRI data, T1-magnetization-prepared rapid gradient-echo (MPRAGE) or fast spoiled gradient echo (FSPGR) and T2-weighted three-dimensional fluid attenuation inversion recovery (3D FLAIR) sequences, ideally with 1 mm^3^ isotropic resolution, underwent a four-step pre-processing: 1. de-identification through the elimination of PHI-containing DICOM headers, 2. DICOM to NIFTI transformation, 3. 6-dof alignment to MNI template orientation, using *ANTS* package ^25^ to first register the T1 image to the 152 MNI template^26^ and then corregister the T2 image to the aligned T1 image. 4. SkullStripping the T1 image using *ROBEX*^*27*^ and using the same stripping mask to SkullStrip the corregistered T2 image. and 5. Correct bias fields in the T1 image using the N4 algorithm from ANTS ^28^.

The volumetric data of different CNS structures were then computed by the LesionTOADS algorithm^29^ implemented in a cloud based medical image-processing platform, QMENTA. LesionTOADS uses an atlas-based technique combining a topological and statistical likelihood atlas for computation of following 12 segmented CNS tissues: Cerebral white matter, Cerebellar white matter, Brainstem, Putamen, Thalamus, Caudate, Cortical gray matter, Cerebellar gray matter, Lesion Volume, Lesion Number, Ventricular CSF and Sulcal CSF.

LesionTOADS results were downloaded from QMENTA server and manually quality checked by an investigator blinded to clinical and functional data (MV). Scans where LesionTOADS segmentation algorithm masks were incorrectly aligned with targeted anatomical structures were excluded from analyses.

### Development and optimization of COMRISv2 models

COMRISv2 models were constructed using random forest (RF) ^30, 31^, a decision-tree-based supervised learning algorithm. A decision tree is a modeling approach that uses multiple features (e.g., MRI volumes) to predict an outcome (e.g., disability) by finding the optimal split (e.g., a specific volume) at each branchpoint in the tree. Tree-based models are prone to “overfit” the data. RF aggregates thousands decision trees and uses a random subset of variables for decision-making at each branchpoint to limit (but not eliminate) the overfit problem. Thus, to further optimize our models, we used the iterative process where the least important variable ranked by variable importance was removed and the RF was rebuilt repeatedly until the root mean square error of the model reached its lowest point. For a visual depiction of this process, see Jackson et al.^32^ and supplemental figure S1. Default tuning parameters (ntree = 500 and mtry= number of variables/3) were used for all models to ensure fair inter-model comparisons.

### Statistical analyses and implemented safeguards to prevent bias

The correlation between observed and predicted outcomes was assessed by Spearman correlation coefficient Rho. The coefficient of determination (R^2^) measuring the proportion of variance of observed outcomes that is explained by the model prediction, as well as the p-value, were calculated from linear regression models. The reproducibility of predicted versus observed outcomes was evaluated by Lin’s concordance correlation coefficient (CCC). The univariate correlations between age, clinical, and MRI outcomes was evaluated using Spearman correlation, the p-value cut-off for significant observations was set to 0.001 to account for 27 pairwise comparisons. All statistical analyses were performed in RStudio Version 1.1.463.

The user-defined privileges in the NDS database assured blinding, while software codification of the algorithms for calculating different scales prevented bias in these calculations. Finally, all models were validated in an independent cohort that did not participate, in any way, in the model development.

## Results

### COMRISv2 model of cognitive disability: SDMT

The COMRISv2 model optimized to predict SDMT score validated in an independent cohort (Rho = 0.674, p-value <0.001, R^2^ = 0.458, CCC = 0.562) (Figure 1A). COMRISv2 SDMT model outperformed the COMRISv1 predictions (Rho = 0.497, p-value <0.001, R^2^ = 0.247, CCC = 0.404) of SDMT score in the same cohort. Age and qMRI features ranked most important in the model, although several semi-qMRI features were also included (Figure 1B).

**Figure 1:**
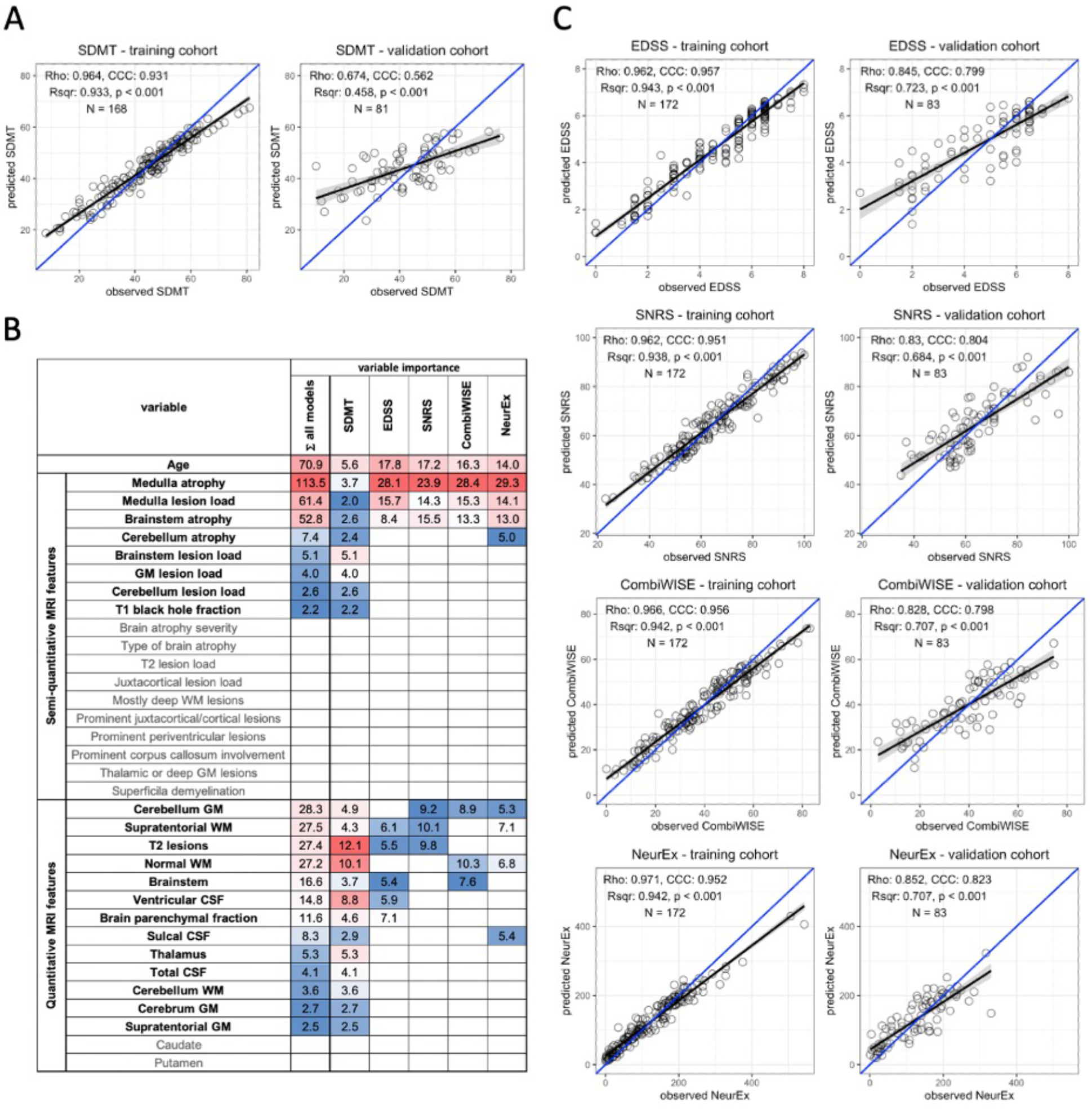
Random forest (RF)-based COMRISv2 models of clinical outcomes. (**A**) RF model of Symbol Digit Modalities Test (SDMT); (**B**) Age, 18 semi-quantitative, and 15 quantitative MRI features were used for RF modelling of the Expanded Disability Status Scale (EDSS), Scripps Neurological Rating scale (SNRS), Combinatorial Weight-adjusted Disability Score (CombiWISE), and digitalized neurological exam score (NeurEx). Variables selected by each model and their respective importance are highlighted here. Variables not selected by any model are depicted in gray. (**C**)RF models of physical disability measured by EDSS, SNRS, CombiWISE, and NeurEx. (**A, B**) The performance of each model was evaluated separately in the training cohort (left plot) and an independent validation cohort (right plot) by plotting observed values on the x-axis and model-predicted values on the y-axis. Spearman Rho, coefficient of determination (Rsqr), p-value (p), Lin’s concordance correlation coefficient (CCC), and number of observations (N) are depicted. Black line represents a fitted linear model with the gray-shaded area corresponding to 95% confidence interval. The blue line represents 1:1 fit corresponding to 100% CCC.

### COMRISv2 models of physical disability: EDSS, SNRS, CombiWISE and NeurEx

COMRISv2 models were also constructed to predict physical disability as measured by four different scales: EDSS, SNRS, CombiWISE, and NeurEx. All models of physical disability performed stronger than the COMRISv2 model for cognitive disability (Figure 1C). The NeurEx scale performed the strongest (Rho = 0.852, p value <0.001, R^2^ = 0.707, CCC = 0.823). Models of physical disability favor semi-qMRI biomarkers reflecting disease burden in the infratentorial compartment (Figure 1B).

### Comparing added value of quantitative volumetric features to COMRISv2 models

Addition of qMRI biomarkers strengthened the performance of the cognitive disability COMRISv2 model. COMRISv2 models for cognitive disability were constructed considering only qMRI measures and only semi-qMRI measures, both in presence and absence of age. Cognitive disability models considering only qMRI measures (Rho = 0.568, p-value <0.001, R^2^ = 0.363, CCC = 0.497) performed slightly better than those considering only semi-qMRI measures (Rho = 0.544, p-value <0.001, R^2^ = 0.282, CCC = 0.474), but the best performance was in the original model considering qMRI, semi-qMRI, and age (Rho = 0.674, p-value <0.001, R^2^ = 0.458, CCC = 0.562).

However, when we performed the same comparison in physical disability models (NeurEx and EDSS), addition of qMRI features did not improve model performance. NeurEx and EDSS models that considered only age and semi-qMRI features outperformed models that included qMRI features (Figure 2).

**Figure 2:**
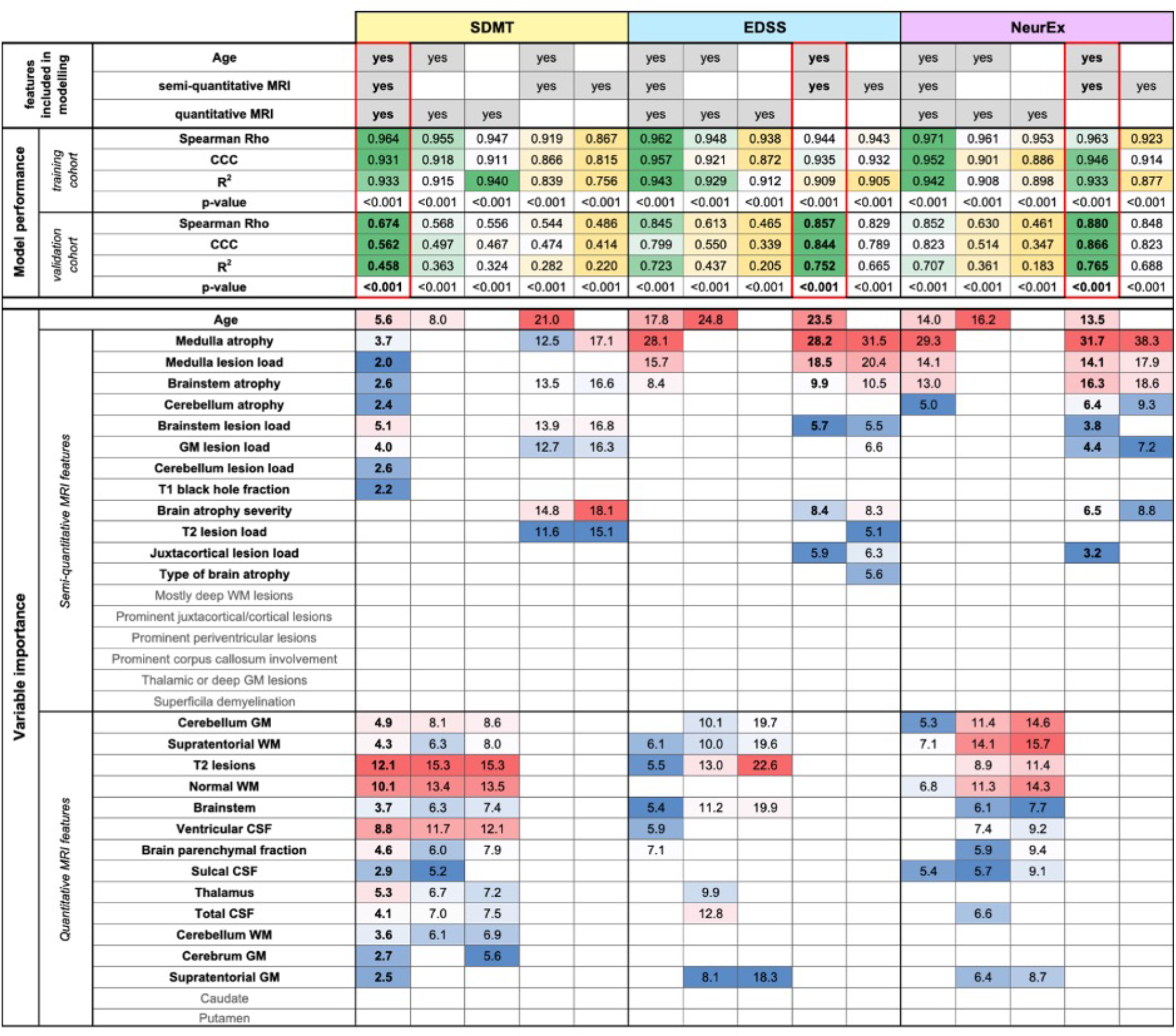
Evaluation of added value of quantitative MRI features. The original random forest (RF) models used Age, semi-quantitative, and quantitative MRI features as an input. We tested how the RF models of Symbol Digit Modalities Test (SDMT), Expanded Disability Status Scale (EDSS), and digitalized neurological exam score (NeurEx) would perform if only semi-quantitative or only quantitative MRI features (with or without age) would perform. The performance of each model was evaluated separately in the training and an independent validation cohort by calculating Spearman Rho, Lin’s concordance correlation coefficient (CCC), coefficient of determination (R2), and p-value. The best performing models are highlighted by red rectangles. Variables selected by each model and their respective importance is also depicted.

### Comparing feature selection between different COMRISv2 models with univariate correlations between MRI biomarkers and clinical outcomes

To facilitate interpretability of COMRIS models, we examined univariate correlations between all MRI features selected by at least one COMRIS model and all clinical outcomes plus age (Figure 3).

**Figure 3:**
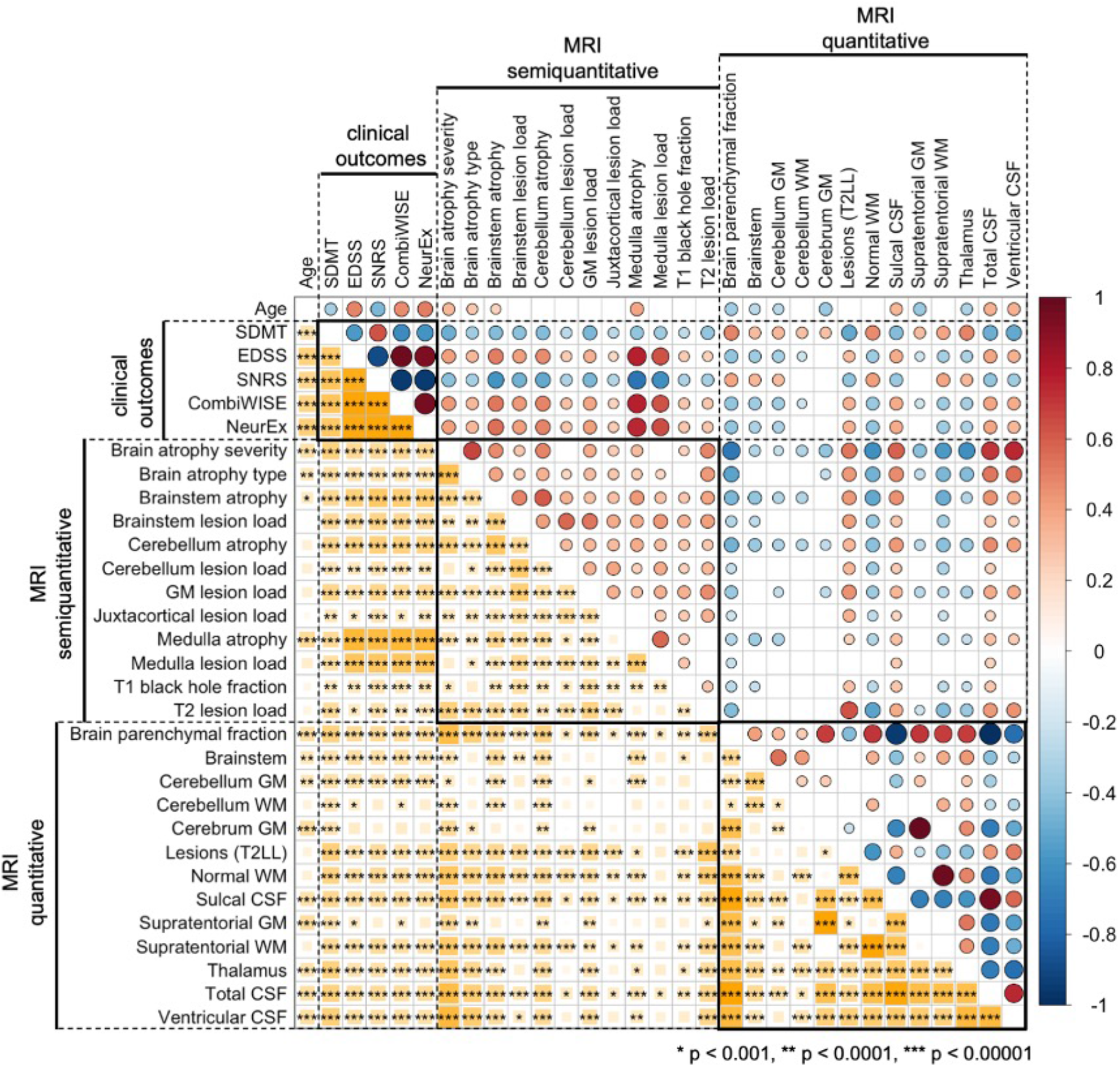
Univariate correlations between age, clinical outcomes, semiquantitative, and quantitative MRI features. Correlations were evaluated using Spearman Rho, with positive correlation in shades of red and negative correlations in shades of blue. The size of the circles above the diagonal and the size of the yellow squares below the diagonal corresponds to the absolute value of Rho. Only correlations with p-value below 0.001 were considered significant (accounting for 31 comparisons performed). The number of stars corresponds to the level of statistical significance.

As it would be expected, all clinical outcomes correlated moderately with age. qMRI outcomes related to CSF and Brain parenchymal fraction (BPF) also correlated with age, although only weakly. From infratentorial structures, only cerebellar gray matter (GM) and semi-qMRI biomarker of brainstem and medulla/upper spinal cord (SC) atrophy correlated with age.

Most qMRI measures correlated with each other, except cerebellar GM, the only infratentorial qMRI biomarker selected by four out of five COMRIS models, which showed only weak correlations with few outcomes. All semi-qMRI biomarkers correlated with each other, but the correlations were generally weak to moderate. Semi-qMRI features also correlated with qMRI features, except medulla/upper SC lesion load that correlated marginally with sulcal CSF. Although medulla/upper SC atrophy correlated with most of qMRI outcomes, these correlations were marginal with qMRI volumetric measures of telencephalon and strongest for brainstem and cerebellum GM volume. Thus, we conclude that qMRI and semi-qMRI measures provide mostly complimentary information, with qMRI outcomes better reflecting telencephalon tissue damage and semi-qMRI outcomes better capturing infratentorial tissue damage.

All clinical outcomes correlated with each other, with SDMT exhibiting only moderate correlations, while other clinical scales correlated strongly. The difference between SDMT and all remaining clinical outcomes was also evident from correlations with MRI outcomes: qMRI biomarkers correlated stronger with SDMT (cognitive disability) in comparison to clinical outcomes that capture predominantly physical disability. In contrast, semi-qMRI biomarkers, especially medulla/upper SC atrophy and lesion load, followed by brainstem atrophy, correlated with non-SDMT clinical outcomes and these correlations were overall stronger than correlations of qMRI measures with SDMT.

## Discussion

With the technological advances that allow reliable measurements of genetic, transcriptomic and proteomic biomarkers in hundreds of patients, data scientists are realizing that the most limiting obstacle in translating these “omics” data into clinically-translatable insights are, surprisingly, poor quality clinical and imaging data. This sentiment is epitomized in the recent review: “It is amazing how bad the standard data sets in the medical domain are (noisy, sparse, wrong, biased etc).” ^33^ Employing unbiased, data-driven approaches to develop more accurate tools for measuring neurological disability and CNS tissue damage and validating both their criterion validity and reproducibility is the way to transcend this conundrum.

*This paper demonstrates the power of ML approach to assemble semi-qMRI and qMRI brain imaging biomarkers into combinatorial models (COMRISv2) that reliably predict neurological disability in MS patients*.

Compared to previously published composite MRI scales, our study has following strengths: 1. The MRI features and their weights are selected using unbiased, data-driven approach; 2. We studied the large cohort of MS patients, with good representation of subjects with progressive MS; 3. COMRISv2 tested both semi-qMRI and qMRI volumetric data; 4. In addition to EDSS, we modeled COMRISv2 predictions of physical disability against highly granular CombiWISE and NeurEx scales, and predictions of cognitive disability against SDMT; 5. Our models are validated in the independent cohort of MS patients that did not contribute, in any way to the development or optimization of the model(s). Literature search identified only one other group that assessed correlation with EDSS in an independent cohort: Bakshi et al observed that their MRDSS1 composite scale showed a trend, but failed to validate correlation with EDSS in new MS cohort (Rho 0.25, p = 0.067, n=55; ^21^). Independent validation is essential to understand true predictive power of composite construct on patients whose data did not contribute to model development. Presented data, congruent with most independent validation studies published, show unequivocally that training cohort results always over-estimate true strength of the relationships.

We also recognize the following limitations of current study: 1. Although our original COMRISv1 computation is publicly available, including detailed guideline for semi-qMRI ratings^22^, and we observed that adherence to those guidelines leads to mean inter-rater variability less than 10%, up till now no external group attempted to reproduce our data. This causes some uncertainty whether other investigators could achieve analogous reproducibility of COMRIS models; 2. We did not test qMRI measures of atrophy or T2LL in the medulla/upper SC, as LesionTOADS algorithm does not provide these outcomes; 3. Our study did not include qMRI biomarkers derived from advanced imaging methods such as MTR or DTI.

While we can’t influence the first limitation, we can address the effect of subsequent two limitations by literature review: First, high quality volumetric SC data require dedicated SC imaging, not available in our patients. Second, published observations suggest that addition of qMRI cervical SC biomarkers would have limited effect on COMRISv2 performance: e.g., the second iteration of Magnetic Resonance Disease Severity Scale (MRDSS2)^21^ demonstrated that addition of upper cervical SC area to MRDSS1 model (which consisted only of brain qMRI features) increased correlation with EDSS from 0.25 to 0.33 (p = 0.013). Both COMRISv1 and COMRISv2 models (using only semi-qMRI features) validated much stronger correlations with EDSS (i.e., Rho = 0.857, p<0.001). Analogously, meta-analysis (21 studies/1933 participants) of dedicated 3T SC imaging showed moderate correlation of cervical SC atrophy with EDSS (Rho=-0.42; p<0.0001).^34^ This likely represents over-estimation, as included studies with small number of participants showed invariably larger correlations. It has been convincingly shown that small studies over-estimate effect size ^35^. Correspondingly, large study (n=1249) published after the afore-mentioned meta-analysis measured Rho -0.315 (p<0.01) for correlation of cervical SC atrophy with EDSS ^36^. These are analogous or smaller univariate correlations as those we observed in COMRIS models between EDSS and two highest ranking semi-qMRI biomarkers: medulla/cervical SC atrophy and T2LL (Figure 3 and ^22^).

Based on the disappointing performance of qMRI compared to semi-qMRI biomarkers in COMRISv2 models, we do not expect that incorporating MTR or DTI data can meaningfully enhance correlations with clinical outcomes for several reasons: 1. These advanced imaging biomarkers have even poorer SNR than volumetric qMRI measures ^6, 10^; and 2. COMRISv2 optimized for CombiWISE or NeurEx already explains close to 70% of physical disability variance in the independent validation cohort, which is exceptionally good performance.

To our knowledge only single study (n=23) included advanced imaging measures (volumetric qMRI measures plus MTR, DTI and MR spectroscopy [MRS] outcomes) to composite MRI models and observed correlations with EDSS ranging from Rho 0.58-0.73 (p = 0.004-0.0001) ^12^. Comparing this (training) cohort with our training cohort results, we conclude that COMRISv2 model (EDSS optimized) already achieved stronger correlation with EDSS (Rho 0.962, p<0.01; Figure 1C). In fact, COMRISv2 outperformed this composite MRI scale with advanced imaging features in its ability to predict EDSS even in the independent validation cohort (Rho 0.845, p<0.001; Figure 1C).

Why is this the case? Intuitively, combining information from structural imaging with biomarkers that may capture cellular/molecular events such as demyelination (MTR/DTI) or neuronal loss (NAA/Cr-Ratio for MRS) should enhance the criterion validity of such construct. Indeed, the cited study supports this intuition as inclusion of the advanced imaging biomarkers enhanced correlation of the models with EDSS in comparison to a single volumetric feature such as T2LL (unfortunately, the authors did not report composite MRI model developed solely from volumetric qMRI features). So why didn’t the inclusion of these advanced imaging measures explain much higher proportion of EDSS variance? One reasons may be the lack of SC biomarkers tested, but we believe that more important, and less appreciated reason is “noise”.

We observed, disappointingly, that addition of volumetric qMRI data enhanced meaningfully only COMRISv2 model of cognitive disability (SDMT). On a contrary, when we forced ML algorithm to derive predictions of physical disability (i.e., CombiWISE and NeurEx) using only semi-qMRI biomarkers, these models achieved stronger correlations with the outcomes in the validation cohort compared to models that contained both qMRI and semi-qMRI features. How can it be? The answer lies in the Figure 2 and Supplementary information: careful examination of the models that contain all features, versus optimized models that discards features with marginal contributions shows invariably better training cohort performance and worse validation for models with all features. This is called “overfit”, caused by the power of the ML algorithm to enhance training model performance beyond criterion validity of contributing biomarkers by using “noise”. “Noise” represents measurements that lack relationship with the process we are trying to model, in our case the MS-associated CNS tissue destruction.

The limitation of the criterion validity of simple volumetric qMRI biomarkers we mentioned in the introduction is exemplified in highly informative postmortem imaging pathological assessment, which showed that SC atrophy (19-24%) strongly under-estimates axonal loss (57-62%) in MS ^37^. Because these imaging data were postmortem, they were not affected by motion artifacts and signal averaging which would further decrease the strength of relationship between imaging biomarker and histologically measured CNS tissue destruction. Therefore, at very best technical imaging conditions the criterion validity of volumetric qMRI biomarkers is limited. Nevertheless, these qMRI biomarkers, especially when measuring large telencephalon structures have validated relationship to CNS tissue destruction in MS: 1. brain atrophy is higher in MS compared to HV; 2. it correlates with disability in large cohorts and 3. It predicts disability progression in long longitudinal studies ^38-40^. Consequently, if we could measure volume of all CNS structures with high accuracy, qMRI biomarkers would likely outperform semi-qMRI biomarkers in all models, as we observed for SDMT version of COMRISv2.

Unfortunately, the test-retest variance (“noise”) of qMRI outcomes increases inversely to the size and contrast of the structure measured and the required scanning time. Additionally, motion artifacts from skeletal muscles, heartbeats, breathing and cerebrospinal fluid pulsation further disadvantage qMRI biomarkers from infratentorial structures. Thus, volumetric biomarkers derived from small infratentorial structures with low MRI contrast from neighboring tissues, that need long acquisition times will have high “noise” (and low SNR)^6^. This explains why infratentorial semi-qMRI biomarkers, while theoretically less sensitive, outperformed infratentorial qMRI features (Figure 1B and Figure 2): because they are measured with higher SNR. The fact that this was not true for semi-qMRI biomarkers of whole brain atrophy or telencephalon T2LL, which ML algorithms found inferior to their qMRI counterparts, strongly support conclusion that the noise in MRI biomarkers measurement in living humans is the deciding limitation of their clinical utility. Research advances to limit this measurement noise may have greater clinical value than development of imaging methods that require longer scanning times and increased complexity of mathematical/physical data manipulations to produce quantitative output.

Generation of qMRI data is expensive and requires specialized skillset. We show that the ability to read clinical brain MRIs in MS neurology practice, when formalized into semi-qMRI outcomes used in COMRIS models, provides validated constructs with surprisingly strong predictive power for clinical and cognitive disability. The CNS tissue damage in the infratentorial compartment, especially in medulla and upper cervical SC is much stronger determinant of physical disability than brain atrophy, or MS lesions in the telencephalon, while the latter two strongly affect cognitive disability.

In conclusion, this study demonstrates remarkable criterion validity of measuring CNS tissue damage in MS by two highly different modalities: neurological examination and brain MRI. There is nothing in clinical data that makes them inevitably poor quality (i.e., noisy, sparse, wrong, biased) if we approach their collection and their aggregation into sensitive and accurate scales with the same scientific rigor used to optimize collection and quantification of omics data. The observations that novel scales of neurological disability with much larger dynamic range than EDSS (i.e., total of 20 possible disability progression steps in the ordinal EDSS scale, versus practically unlimited range of CombiWISE [0-100 continuous scale] and NeurEx [0 to theoretical maximum of 1349] values) validated comparably, or even outperformed EDSS demonstrates that implementing data-driven approached to development of new clinical scales allows increasing sensitivity without limiting their accuracy. The CCC of 0.866 between semi-qMRI features-derived COMRISv2 and NeurEx in the independent validation cohort indicates that scientists have a reliable inexpensive tool at their fingertips that can predict the most granular scale of neurological disability we currently have in MS research.

## Data Availability

All data is available upon request.

## Acknowledgments

We would like to thank our clinical team (Dr. Alison Wichman, Ms. Michelle Woodland, and Ms. Tiffany Hauser), patients, and caregivers at the Neuroimmunological Diseases Section for partnering with us in this project. Funding for this study was provided by the Division of Intramural Research, National Institute of Allergy and Infectious Diseases, National Institutes of Health.

## Author contributions statement

B.B. designed the project and guided all aspects of data collection and analyses. M.S. performed neurological exam and collected clinical data. V.P. and D.M-D. implemented LesionTOADS algorithm into QMENTA platform. E.K., M.V., P.K. and B.B. wrote the first draft of the paper P.K. prepared figures and all authors reviewed and edited the draft for intellectual content.

## Additional Information

E.K., M.V., P.K., M.S., and B.B. declare no competing interests. V.P. and D.M-D. were employees of Qmenta at the time of the study.

